# Linking rattiness, geography and environmental degradation to spillover *Leptospira* infections in marginalised urban settings: an eco-epidemiological community-based cohort study in Brazil

**DOI:** 10.1101/2021.09.21.21263884

**Authors:** Max T. Eyre, Fábio N. Souza, Ticiana S. A. Carvalho-Pereira, Nivison Nery, Daiana de Oliveira, Jaqueline S Cruz, Gielson A. Sacramento, Hussein Khalil, Elsio A. Wunder, Kathryn P. Hacker, James E. Childs, Mitermayer G. Reis, Mike Begon, Peter J. Diggle, Albert I. Ko, Emanuele Giorgi, Federico Costa

**Affiliations:** Centre for Health Informatics, Computing, and Statistics, Lancaster University Medical School, UK; Liverpool School of Tropical Medicine, Liverpool, UK; Institute of Collective Health, Federal University of Bahia, Salvador, Bahia, Brazil; Swedish University of Agricultural Sciences, Umeå, Sweden; Oswaldo Cruz Foundation, Brazilian Ministry of Health, Salvador, Bahia, Brazil; Department of Epidemiology of Microbial Diseases, Yale School of Public Health, New Haven, USA; University of Pennsylvania, Philadelphia, Pennsylvania, USA; Department of Evolution, Ecology and Behaviour, University of Liverpool, Liverpool, UK

**Keywords:** zoonotic spillover, leptospirosis epidemiology, transmission pathways, rat reservoir, rodent-borne zoonoses, multivariate model-based geostatistics, joint spatial modelling

## Abstract

**Background:** Zoonotic spillover from animal reservoirs is responsible for a significant global public health burden, but the processes that promote spillover events are poorly understood in complex urban settings. Endemic transmission of *Leptospira*, the agent of leptospirosis, in marginalised urban communities occurs through human exposure to an environment contaminated by bacteria shed in the urine of the rat reservoir. However, it is unclear to what extent transmission is driven by variation in the distribution of rats or by the dispersal of bacteria in rainwater runoff and overflow from open sewer systems.

**Methods:** We conducted an eco-epidemiological study in a high-risk community in Salvador, Brazil, by prospectively following a cohort of 1,401 residents to ascertain serological evidence for leptospiral infections. A concurrent rat ecology study was used to collect information on the fine-scale spatial distribution of ‘rattiness’, our proxy for rat abundance and exposure of interest. We developed and applied a novel geostatistical framework for joint spatial modelling of multiple indices of disease reservoir abundance and human infection risk.

**Results:** The estimated infection rate was 51.4 (95%CI 40.4, 64.2) infections per 1,000 follow-up events. Infection risk increased with age until 30 years of age and was associated with male gender. Rattiness was positively associated with infection risk for residents across the entire study area, but this effect was stronger in higher elevation areas (OR 3.27 95%CI 1.68, 19.07) than in lower elevation areas (OR 1.14 95%CI 1.05, 1.53).

**Conclusions:** These findings suggest that, while frequent flooding events may disperse bacteria in regions of low elevation, environmental risk in higher elevation areas is more localised and directly driven by the distribution of local rat populations. The modelling framework developed may have broad applications in delineating complex animal-environment-human interactions during zoonotic spillover and identifying opportunities for public health intervention.

**Funding:** This work was supported by the Oswaldo Cruz Foundation and Secretariat of Health Surveillance, Brazilian Ministry of Health, the National Institutes of Health of the United States (grant numbers F31 AI114245, R01 AI052473, U01 AI088752, R01 TW009504 and R25 TW009338); the Wellcome Trust (102330/Z/13/Z), and by the Fundacção de Amparo à Pesquisa do Estado da Bahia (FAPESB/JCB0020/2016). M.T.E is supported by an MRC doctorate studentship. F.N.S. participated in this study under a FAPESB doctorate scholarship.

## 1 Introduction

Zoonotic spillover, the transmission of pathogens from infected vertebrate animals to humans, is responsible for a significant public health burden globally. Understanding the processes that promote spillover transmission is essential for improving our ability to predict and prevent spillover events, but for many zoonoses, such as *Leptospira interrogans, Escherichia coli* O157 and *Giardia* spp., they are poorly understood [1]. This is due to the complex nature of the spillover system, in which the probability of transmission is governed by dynamic interactions in space and time between ecological, epidemiological, behavioural and immunological factors that determine pathogen pressure, exposure and host susceptibility. Zoonotic spillover research must explore interactions between the environment, disease reservoirs and local epidemiology, presenting two central challenges: i) the need for transdisciplinary studies at the animal-human disease interface (a One Health approach) that accurately collect data on multiple components of the spillover process at common temporal and spatial scales at which these events take place; ii) the development of integrative approaches to jointly analysing these diverse datasets within a spatially and temporally explicit framework [1–3].

Leptospirosis, a neglected zoonotic disease caused by pathogenic bacteria from the genus *Leptospira*, is an important example of zoonotic spillover. Globally, it is estimated to cause more than one million cases and over 58,000 deaths each year [4], with an annual global burden of 2.9 million DALYs [5]. This burden falls heavily on marginalised urban populations in low- and middle-income countries who live in areas characterised by high population density, poor quality housing and inadequate provision of healthcare, sanitation, and waste management services. In these settings, leptospiral infection occurs through contact with water or soil contaminated with leptospires shed in the urine of the principal reservoir, the Norway rat (*Rattus norvegicus*) [6]. These areas produce the socio-ecological conditions that allow rodent populations to proliferate and leptospires to persist for long periods in the environment [7]. Residents consequently have frequent, intense and largely unavoidable exposure to the contaminated environment, often exacerbated by their geographical vulnerability to flooding events [8]. In response, the World Health Organisation (WHO) has convened the Leptospirosis Burden Epidemiology Reference Group (LERG) which has recommended “Targeted intervention based on the improved knowledge of disease ecology” [9], highlighting the current knowledge gap for *Leptospira* transmission mechanisms and target points for effective intervention.

Multiple studies have helped to elucidate key aspects of the *Leptospira* transmission cycle in urban settings, identifying socioeconomic vulnerability, household environment and behavioural exposures as important determinants of infection risk [7, 10–19]. However, these variables have been unable to explain fine-scale spatial variation in risk [10, 12]. This is likely to be driven by the high spatial and temporal heterogeneity in environmental risk, observed in recent studies of *Leptospira* in soil, and surface and sewage waters [6, 20, 21]. These findings lead to two key questions: i) to what extent does environmental contamination by localised rat shedding drive infection risk, rather than exposure to leptospires that have been dispersed by rainwater runoff and overflowing sewer systems; and ii) how does this change across the geography of a community, for example at different elevation levels?

Establishing a dynamic link between rats, the environment and *Leptospira* transmission is complicated by the difficulty of measuring and modelling the rat contamination process. However, urban Norway rats have been found to have high *Leptospira* prevalence and shedding rates worldwide [22–28]. This suggests that rat abundance may be predictive of environmental risk, and could be used as a proxy for this shedding process. While several studies have identified associations between infection risk and household rat sightings and infestation [10, 12, 22, 29–31], their ability to explore fine-scale spatial variation in risk was limited by a reliance on household infestation surveys or aggregation of incidence and abundance indices to a common coarse spatial scale. All modelled abundance as a regression covariate, thereby not accounting for uncertainty in its measurement. The absence of methods to formally integrate abundance into analyses of spillover mechanisms is an issue for rodent-borne zoonoses more widely [3, 32].

There is no gold-standard index of abundance and field teams use a range of imperfect indices, such as traps, infestation surveys and track plates. In our previous work, we developed a multivariate generalized linear geostatistical model for joint spatial modelling of multiple imperfect abundance indices [33]. We use the term ‘abundance’ loosely here to denote all ecological processes that are associated with animal abundance, for example animal presence and activity, and that can be used to quantify exposure to a disease of interest. This methodology was then used to model the spatial distribution of ‘rattiness’, our proxy for rat abundance, at a fine scale within a community in Salvador, Brazil [33]. The spatial distribution of rattiness was highly heterogeneous, suggesting that it could be a driver of micro-heterogeneity in infection risk.

To analyse reservoir host abundance and infection data at fine spatial scales, we propose that a framework should i) account for spatial correlation in human and reservoir host data; ii) jointly model multiple imperfect indices of abundance while accounting for the appropriate sampling distribution of each index; iii) account for uncertainty in abundance indices, iv) allow for the prediction of abundance and infection risk at all locations within the study area, and v) quantify the uncertainty associated with those predictions. Several studies have attempted to model spatial associations between disease reservoir or vector abundance and human infection for leptospirosis [34–36], tularemia [37]; Lyme disease [38]; West Nile Virus [39]; dengue fever [40]; and Lassa fever [41]. However, none of the approaches used satisfy all five of the above conditions. The development of new tools for the joint spatial analysis of abundance and human infection may consequently be beneficial for the study of other zoonoses and vector-borne diseases [42].

The aim of this study was to develop a flexible modelling framework for zoonotic spillover to explore whether rattiness, acting as a proxy for local leptospiral contamination by Norway rats, can explain spatial heterogeneity in leptospiral transmission in a high-risk urban community in Brazil. We extend the rattiness framework of Eyre et al. [33] to include human infection risk. We describe findings from a transdisciplinary eco-epidemiological study which comprises a prospective community-based cohort study with two serosurveys and a fine-scale rat ecology study. The ecology study was used to collect information on the spatial distribution of rat abundance, our exposure of interest, in the period between the two surveys using multiple abundance indices. Then, we explore associations between infection risk, rattiness and a range of measured environmental and individual risk factors.

## 2 Materials & methods

### 2.1 Study design

#### 2.1.1 Study area

The study was conducted in Pau da Lima community (13°32’53.47” S; 38°43’51.10” W), a marginalised informal settlement located in the city of Salvador, Northeast Brazil. The study site has an area of 0.25*km*^2^ and is characterised by three connected valleys with large elevation gradients, high population density and a heterogeneous environment of vegetation, paved surfaces and exposed soil (Figure 1). There are significant gradients in socioeconomic status and infrastructure quality over small elevation increases - with the most marginalised members of the community living at lower elevations. The community suffers from low quality housing, poor provision of waste management services and inadequate drainage and sanitation systems [12, 43]. Residents are consequently often unable to avoid intense exposure with mud and floodwater. These factors result in abundant rat populations [33] and a high estimated annual *Leptospira* infection rate of 35.4 (95% CI, 30.7, 40.6) infections per 1,000 annual follow-up events [12]. For this reason, Pau da Lima has become an exemplar for investigating urban *Leptospira* transmission in Brazil over the last 15 years.

**Figure 1:**
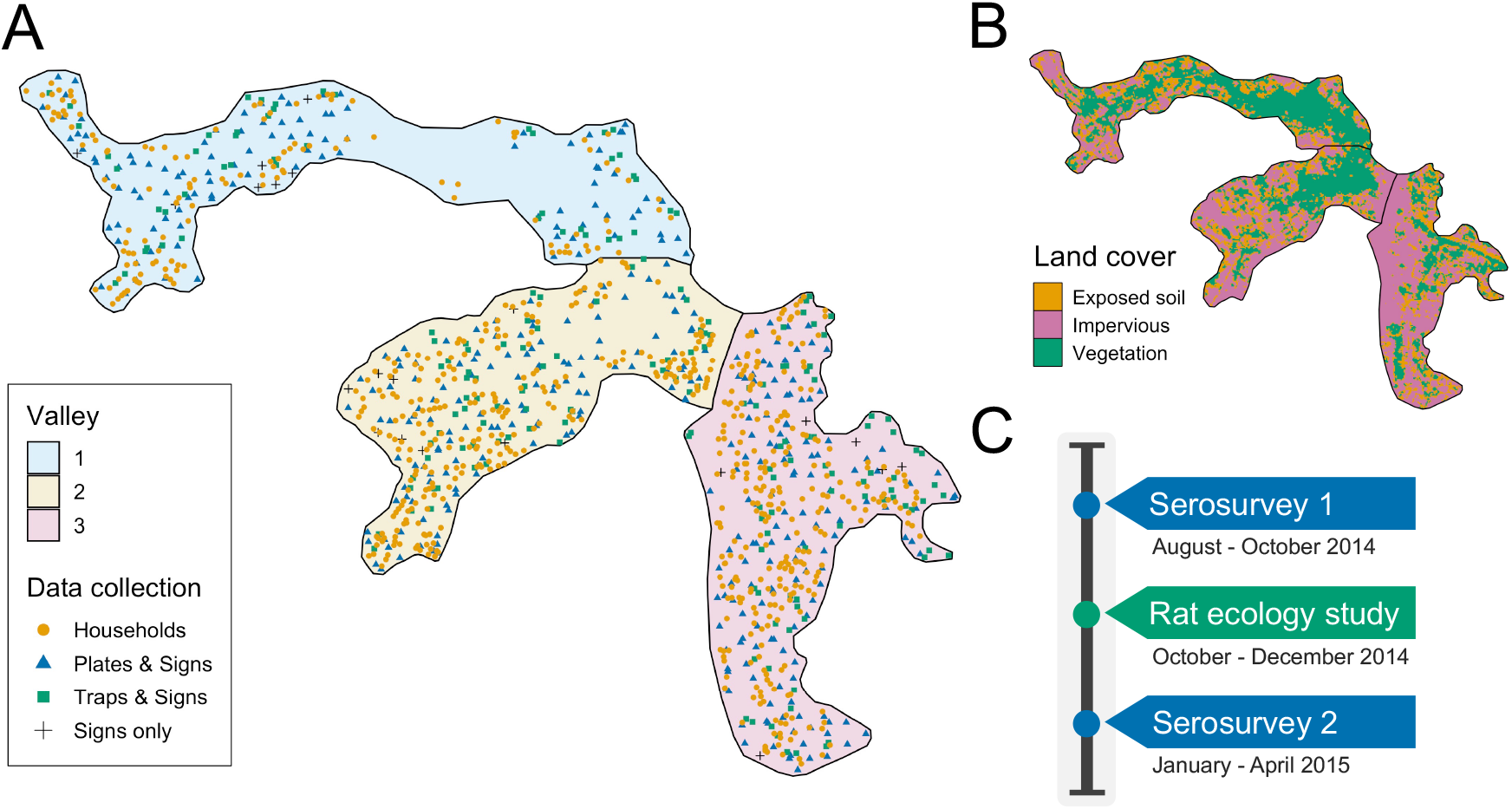
A) Map of the three valleys within the study site in Pau da Lima, with household locations for the serosurveys marked as orange circles. Locations sampled in the the rat ecology study are shown for each of the rat abundance indices as follows: Plates & Signs (track plates, burrows, faeces and trails), Traps & Signs (traps, burrows, faeces and trails) and Signs only (burrows, faeces and trails); B) Land cover classification map (impervious cover is defined as man-made structures e.g. pavement and buildings); C) Study timeline for the two community serosurveys and rat ecology study.

#### 2.1.2 Serosurveys

We conducted a prospective community cohort study with two serosurveys carried out in August-October 2014 and January-April 2015. After an initial census of the study site, all ground floor households were visited and inhabitants who met the eligibility criteria of ≥ 5 years of age who had slept ≥ 3 nights in the previous week in a study household were invited to join the study.

During each survey trained phlebotomists collected blood samples from participants and administered a modified version of the standardised questionnaire used previously [12, 29]. Information was collected on demographic and socioeconomic indicators, household environmental characteristics and exposures to potential sources of environmental contamination in the previous six months (the average time between the two serosurveys). Study data were collected and managed using REDCap electronic data capture tools [44] and all individual data were anonymised. The locations of sampled households are shown in Figure 1A. If an individual was not found during a sample collection visit their house was revisited at least five times on different days of the week.

The microscopic agglutination test (MAT) was used to determine titers of agglutinating antibodies against pathogenic *Leptospira* in sera obtained from the blood samples collected in each serosurvey. Serological samples were reacted with a panel of two *Leptospira* reference strains that are dominant in Pau da Lima: *Leptospira interrogans* serovars Copenhageni (COPL1) and Cynopteri 3522C (C3522C). These two strains have been shown to have the same performance in identifying MAT seroconversion in our prospective studies as the WHO recommended battery of 19 reference serovars. When agglutination was observed at a dilution of 1:50, the sample was titrated in serial two-fold dilutions to determine the highest agglutination titer. The study outcome of leptospiral infection was defined as seroconversion, an MAT titer increase from negative to ≥ 1:50, or a four-fold increase in titer for either serovar between paired samples from cohort subjects. All laboratory analyses were performed in the Laboratory Pathology and Molecular Biology at Fiocruz, Salvador. As part of quality control procedures two independent evaluations were conducted by Yale University for all infected subjects and 8% of all samples, with high concordance between results.

#### 2.1.3 Rat ecology study

To estimate exposure risk due to local rat contamination between the two serosurveys, a cross-sectional rat ecology study was conducted from October to December 2014. As has been described previously [33], the aim of this study was to collect data on the fine-scale spatial variation in rat reservoir population abundance. Data were collected for five indices of rat abundance: live trapping, track plates, number of active burrows present, presence of faecal droppings and presence of trails. Rat trapping was carried out at 189 locations, randomly distributed across the study area (see Panti-May et al. [45]). Two traps were deployed for 4 consecutive 24-hour trapping periods at each location. Trapping success and trap closure without a rat, a common malfunction, were recorded after each 24-hour period. Track plates were placed at 415 locations for two consecutive 24-hour periods following the standardised protocol for placement and survey developed and validated previously [46], with five plates placed at each location in the shape of a ‘five’ on a die. After each 24-hour period plates were repainted and any lost plates were recorded and replaced. On the first day of trapping or plate placement, a survey for signs of rat infestation, adapted from the Centers for Disease Control and Prevention [47] and validated in the study area [29], was conducted within an area of 10m radius around each trapping or plate location to record the number of active burrows and the presence of faecal droppings and trails. In total, 595 independent locations were sampled for traps, track plates and the three survey indices for signs of rat infestation. The spatial distribution of these locations is shown in Figure 1A. At 21 locations, theft and local gang violence meant that data for track plates and traps was not collected and only the three survey indices for signs of rat infestation were used.

#### 2.1.4 Environmental data

In addition to the environmental survey conducted at each household location, we also collected information for three spatially continuous environmental variables: elevation relative to the bottom of each valley, distance to large public refuse piles and the proportion of land cover classified as impervious (man-made structures) within a 30-metre radius. The land cover variable was created from Digital Globe’s WorldView-2 satellite imagery (8 bands) taken on February 17, 2013 which was classified using a maximum likelihood supervised algorithm and validated with ground truthed data collected from 20 randomly selected sites of size 5m by 5m. The classification map is shown in Figure 1B.

#### 2.1.5 Ethics

Participants were enrolled according to written informed consent procedures approved by the Institutional Review Boards of the Oswaldo Cruz Foundation and Brazilian National Commission for Ethics in Research, Brazilian Ministry of Health (CAAE: 01877912.8.0000.0040) and Yale University School of Public Health (HIC 1006006956).

For the rat ecology study, the ethics committee for the use of animals from the Oswaldo Cruz Foundation, Salvador, Brazil, approved the protocols used (protocol number 003/2012), which adhered to the guidelines of the American Society of Mammalogists for the use of wild mammals in research [48] and the guidelines of the American Veterinary Medical Association for the euthanasia of animals [49]. These protocols were also approved by the Yale University’s Institutional Animal Care and Use Committee (IACUC), New Haven, Connecticut (protocol number 2012–11498).

### 2.2 Joint modelling rat abundance and human infection: the rattiness-infection framework

The developed geostatistical modelling framework jointly models multiple rat abundance indices as measurements of a common latent process, called rattiness. Rattiness at each household location contributes to the risk of infection for all inhabitants, in addition to other measured individual or household-level explanatory variables.

We model the rat abundance data following a similar structure to that previously outlined [33]. Let *R*(*x*) denote a spatially continuous stochastic process, representing rattiness. The rat data then consist of a set of outcomes *Y*_*i*_ = (*Y*_*i,k*_ : *k* = 1, …, 5), for *i* = 1, …, *N*_*r*_, collected at a discrete set of locations *X* = {*x*_*i*_ : *i* = 1, …, *N*_*r*_}. The outcome variables *Y*_*k*_ : *k* = 1, …, 5 are the set of five rat abundance indices that provide information about *R*(*x*): traps (*k* = 1), track plates (*k* = 2), number of burrows (*k* = 3), presence of faecal droppings (*k* = 4) and presence of trails (*k* = 5).

Human data are collected from *N*_*h*_ households and consist of an infection outcome *Z*_*i,j*_ for individual *j* at household location *i*, for *i* = *N*_*r*_ + 1, …, *N*_*r*_ + *N*_*h*_, collected at a discrete set of locations *X* = {*x*_*i*_ : *i* = *N*_*r*_ + 1, …, *N*_*r*_ + *N*_*h*_}.

Let “[*·*]” be a shorthand notation for “the probability distribution of *·*.” We write 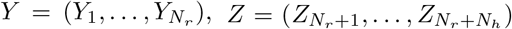 and 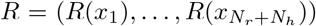. We assume that the *Y*_*i,k*_ : *k* = 1, …, 5 and *Z*_*i,j*_ are conditionally independent given *R*(*x*_*i*_), from which it follows that

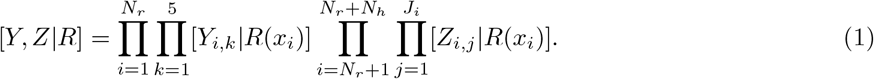

where [*·*] is a shorthand notation for “the distribution of” and *J*_*i*_ denotes the number of individuals at household *i*. This model structure is shown schematically in Figure 2.

**Figure 2:**
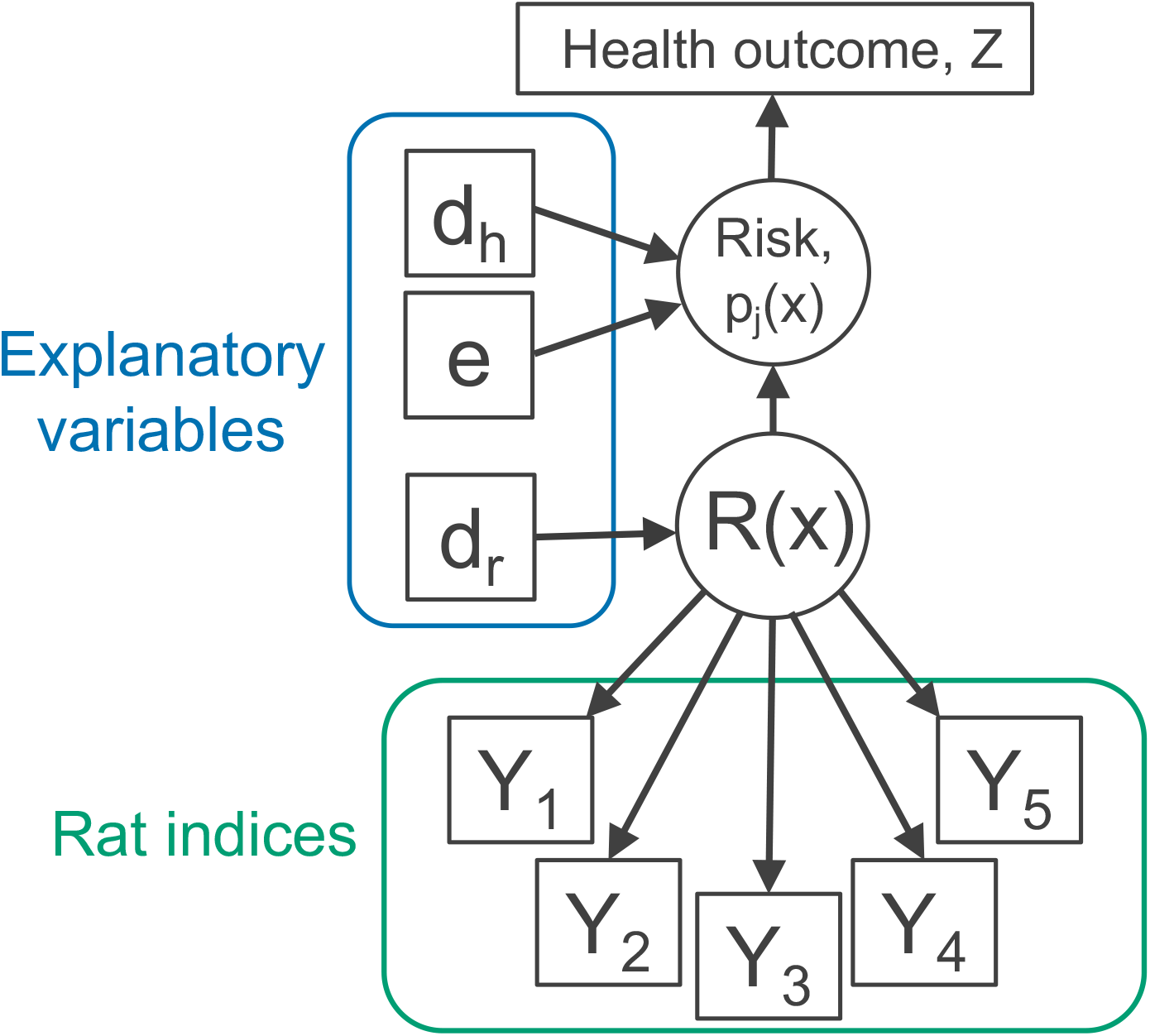
Directed acyclic graph (DAG) of the rattiness-infection model framework. *R*(*x*) is the value of a spatially continuous stochastic rattiness process at location *x*. The outcome variables *Y*_*k*_ : *k* = 1, …, 5 are the set of five rat abundance indices that provide information about *R*(*x*): traps (*k* = 1), track plates (*k* = 2), number of burrows (*k* = 3), presence of faecal droppings (*k* = 4) and presence of trails (*k* = 5). The outcome variable *Z*_*i,j*_ is the observed health outcome, in this case this represents infection status. The terms *d*_*h*_ and *d*_*r*_ represent the sets of spatially continuous explanatory variables which contribute to spatial variation in infection risk in humans and *R*(*x*), respectively. The terms *d*_*h*_ and *d*_*r*_ are not mutually exclusive groups of explanatory and the same variables may contribute to both infection risk and *R*(*x*). The term *e* represents a set of individual- and household-level explanatory variables which contribute to variation in infection risk. Square objects correspond to observable variables, and circles to latent random variables.

#### 2.2.1 Rattiness

We define rattiness at location *x* as

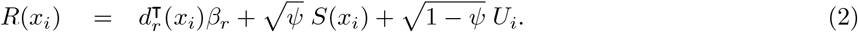

The terms on the right-hand side of 2 have the following interpretations: *d*_*r*_(*x*_*i*_) is a vector of explanatory variables with associated regression coefficients *β*_*r*_; *U*_*i*_ is a set of independently and identically distributed zero-mean Gaussian variables with unit variance; *S*(*x*_*i*_) is a stationary and isotropic spatial Gaussian process; *ψ* ∈ (0, 1) regulates the relative contributions of spatially structured variation, *S*(*x*_*i*_), and unstructured random variation, *U*_*i*_, to *R*(*x*_*i*_).

For the Gaussian process, *S*(*x*_*i*_), we specify an exponential spatial correlation function:

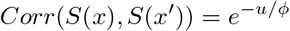

where *u* = ||*x* − *x*′|| is the Euclidean distance between *x* and *x*′, and *ϕ* regulates how fast the spatial correlation decays to zero with increasing distance u.

#### 2.2.2 Rat abundance outcomes

The variable *Y*_*i*,1_, conditionally on *R*(*x*_*i*_), is a binomial variable representing the number of traps, out of *n*_*i*,1_, in which rats were captured. We assume that the times of rat captures from a trap follow a time-varying inhomogeneous Poisson process with intensity *t*_*i*_*μ*_2_(*x*_*i*_), where *t*_*i*_ is the time (in days) for which a trap is operative and log{*μ*_2_(*x*_*i*_)} = *α*_1_ + *σ*_1_*R*(*x*_*i*_). It follows that the probability of capturing a rat is

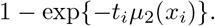

If a trap is found closed without a rat, we assume that the trap was disturbed and set *t* = 0.5. In all other cases, *t* = 1 day.

*Y*_*i*,2_, is the number of track-plates, out of *n*_*i*,2_, that show presence of rats. We model this as a binomial variable with *n*_*i*,2_ trials and probability *μ*_2_(*x*_*i*_) where log{*μ*_2_(*x*_*i*_)*/*(1 − *μ*_2_(*x*_*i*_)} = *α*_2_ + *σ*_2_*R*(*x*_*i*_).

*Y*_*i*,3_, is the number of active rat burrows found at location *x*_*i*_. We model this as a Poisson variable with rate *μ*_3_(*x*_*i*_) where log{*μ*_3_(*x*_*i*_)} = *α*_3_ + *σ*_3_*R*(*x*_*i*_).

The variables *Y*_*i*,4_ and *Y*_*i*,5_ are binary indicators taking value 1, if at least one faecal dropping or trail, respectively, was found at location *x*_*i*_ and 0 otherwise. We model the probability of finding a sign of faecal droppings or trails, *μ*_4_(*x*_*i*_) and *μ*_5_(*x*_*i*_), using logit-linear regressions log{*μ*_4_(*x*_*i*_)*/*(1 − *μ*_4_(*x*_*i*_))} = *α*_4_ + *σ*_4_*R*(*x*_*i*_) and log{*μ*_5_(*x*_*i*_)*/*(1 − *μ*_5_(*x*_*i*_))} = *α*_5_ + *σ*_5_*R*(*x*_*i*_).

#### 2.2.3 Human infection outcome

Conditionally on *R*(*x*_*i*_), we model the binary human infection outcome *Z*_*i,j*_ as a Bernoulli variable with the probability, *p*_*j*_(*x*_*i*_), that individual *j* at location *i* is infected. This is modelled with a logit link function and the following linear predictor

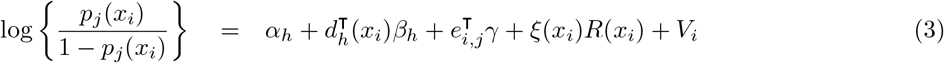

where: *d*_*h*_(*x*_*i*_) is a vector of spatially continuous explanatory variables with associated regression coefficients *β*_*h*_; *e*_*i,j*_ is a vector of household-level and individual-level explanatory variables with associated regression coefficients *γ*; *V*_*i*_ is a set of independently and identically distributed zero-mean Gaussian variables with variance *σ*^2^ representing unexplained household-level variation; *ξ*(*x*_*i*_) regulates the contribution of rattiness to risk of infection.

### 2.2.4 Parameterising to test for an interaction with elevation

To explore variation in the role of local rat populations in transmission within different sections of the study area, *ξ* was parameterised to test for an interaction between rattiness and household elevation level on human infection risk. This was implemented by dividing the study area into three elevation levels with an equal number of households in each: low (0 − 6.7*m* from bottom of valley), medium (6.7 − 15.6*m*) and high (*>* 15.6*m*). We then define the set of household locations in each low, medium and high elevation level as *x*_*low*_, *x*_*med*_ and *x*_*high*_, respectively. Three values of *ξ* were then estimated such that:

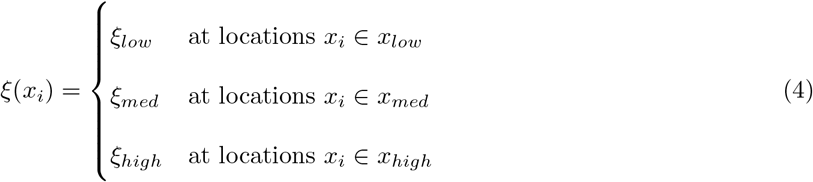

### 2.3 Variable selection

#### 2.3.1 Predictors of rattiness

The exploratory analysis for the rattiness model followed the steps developed and described previously [33]. Firstly, we explored the functional form of the relationship between rattiness and three continuous explanatory variables: relative elevation, distance to large refuse piles and land cover type. To do this, we fitted a simplified rattiness model that did not include covariates or account for spatial correlation. Rattiness is consequently modelled purely as unstructured random variation; hence *R*(*x*_*i*_) = *U*_*i*_ [33]. We then computed the predictive expectation of this simplified rattiness process, *Û*_*i*_, at all locations for which rat index measurements were observed. A generalized additive model (GAM) [50] was then fitted to the *Û*_*i*_ with the three explanatory variables and the shape of each fitted smooth function was used to assess whether the relationship between each variable and rattiness was linear. Non-linear relationships were modelled using linear splines based on the identified functional form, with knots placed at relative elevations of 8m and 22m, and at a distance from large refuse piles of 50m (see Supplementary file 1). For variable selection, linear models with all combinations of these variables were fitted and ranked by their Akaike Information Criterion (AIC) value [51]. The model with the lowest AIC included all of the variables and their linear splines (Supplementary file 2).

Following the methodology outlined previously [33], we fitted the full geostatistical rattiness model using the variables selected in Section 2.3.1. We then plugged in the maximum likelihood estimates and made predictions for rattiness at all human household locations; here, the predictive target is 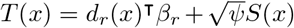 rather than *R*(*x*) as defined by (2) because the predicted value of the spatially uncorrelated *U*(*x*) at any location *x* where rat abundance indices have not been recorded is zero. The expectation of this predictive distribution was then computed to provide an estimate of mean predicted rattiness at all household locations. This was then used as an exploratory covariate in the following section.

#### 2.3.2 Risk factors for human infection

All explanatory variables were grouped into the following four domains: social status, household environment, occupational exposures and behavioural exposures. A group of *a priori* confounding variables was then identified, with age, gender and household per capita income selected based on previous findings [10–12], and valley also included to account for otherwise unmeasured differences between the three valley regions within the study area.

The relationship between continuous explanatory variables and infection risk (on the log-odds scale) was assessed for linearity by fitting a GAM while controlling for the four confounders. As before, non-linear relationships were modelled using linear splines based on the identified functional form. Age was modelled with a knot at 30 years old, education at 5 years and relative elevation at 20m (Supplementary file 1). A univariable analysis was conducted to explore the relationship between each explanatory variable and infection risk while controlling for the four *a priori* confounding variables. Crude and adjusted odds ratios were estimated using a mixed effects logistic regression with a random effect to account for unexplained variation at the household-level.

For the multivariable model, variable selection was conducted within each domain separately. Mixed-effect logistic regression models were fitted for all combinations of the variables in each domain and were ranked by their Akaike Information Criterion (AIC) value (Supplementary file 2). Variables in the model with the minimum AIC value were selected for each domain. Age, gender, household per capita income and valley were controlled for in all models throughout this process. Then, the variables selected from each domain were combined and the mean predicted rattiness estimate (obtained in Section 2.3.1 at each household location) was included with an interaction with elevation level. This set of variables was reduced once more following the same process and all selected variables were included in the final multivariable model (Supplementary file 3).

### 2.4 Model fitting

All rat and human variables selected in Section 2.3 were then included in the full joint model defined in Equations (2) and (3). We fit this model using the Monte Carlo maximum likelihood (MCML) method [52] as described in Supplementary file 4, and compute 95% confidence intervals by re-fitting the model for 1,000 parametric bootstraps.

### 2.5 Prediction maps

The maximum likelihood parameter estimates were then used to make prediction maps for rattiness and infection risk as follows.

To map a general predictive target, *T* (*x*) say, we first define 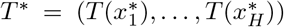, where 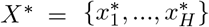 is a finely spaced grid of locations to cover the region of interest. We then draw samples from the predictive distribution of *T*\*, i.e. its conditional distribution given all relevant data. These samples can then be used to compute any desired summary of the predictive distribution. In our analysis, we used as summaries the expectation and 95% prediction interval.

Our first predictive target is rattiness, for which 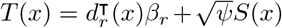. Our second is human infection risk, for which 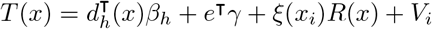. In either case, we first sample from [*R*|*W* ; *θ, ω*] using the same sampling algorithm as for maximizing the likelihood in Section 2.2, with the parameters *θ* and *ω* fixed at their maximum likelihood estimates. After obtaining samples *r*_(*b*)_, *b* = 1, …, *B*, we then sample from [*T* *|*r*_(*b*)_], which in both cases follows a multivariate Gaussian distribution with mean and covariance matrix easily obtained from their joint Gaussian distribution, [*R, T*\*]. The resulting values, 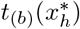, *h* = 1, …, *H*; *b* = 1, …, *B* constitute *b* samples drawn from [*T* *|*W*] as required. Note that each 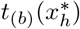, *h* = 1, …, *H* is a sample from the joint predictive distribution of the complete surface of *T*(*x*) over the whole of the region of interest and can therefore be used to make inferences about spatially aggregated properties of *T*(*x*) if required.

### 2.6 Data accessibility

Data and code used in this analysis are publicly available at https://github.com/maxeyre/Rattiness-inf ection-framework and have been published [53]. However, household coordinates and valley ID have been removed from the human data to ensure participant anonymity. The analysis was conducted using R [54] and the following packages: tidyverse [55], mgcv [56], PrevMap [57], MuMIn [58] and lme4 [59].

## 3 Results

### 3.1 Study overview

In Pau da Lima, we identified 3,179 eligible residents using a baseline community census, household visits and through other members of the household. Of these, 2,018 (63.4%) individuals consented to join the study and provided a blood sample in the first serosurvey (August-October 2014). As a result of loss to follow-up, only 1,401 (69.4%) of these participants (from 669 households) completed the second serosurvey (January-April 2015). Individuals were lost to follow-up because they could not be found after at least five attempts (44.4%), had moved out of the study area (31.1%) or did not wish to provide a second blood sample (19.8%). An overview of participant recruitment is provided in Figure 3. Individuals lost to follow-up were similar in age to those who remained in the study cohort (mean 29.0 and 28.8 years old, respectively, t-value= − 0.37, *df* = 1288.5, *p* = 0.7) but were more likely to be male (49.8% male compared to 42.6%, χ^2^ = 8.5, *df* = 1, *p<* 0.01). A full description of the study cohort is included in Supplementary file 5.

**Figure 3:**
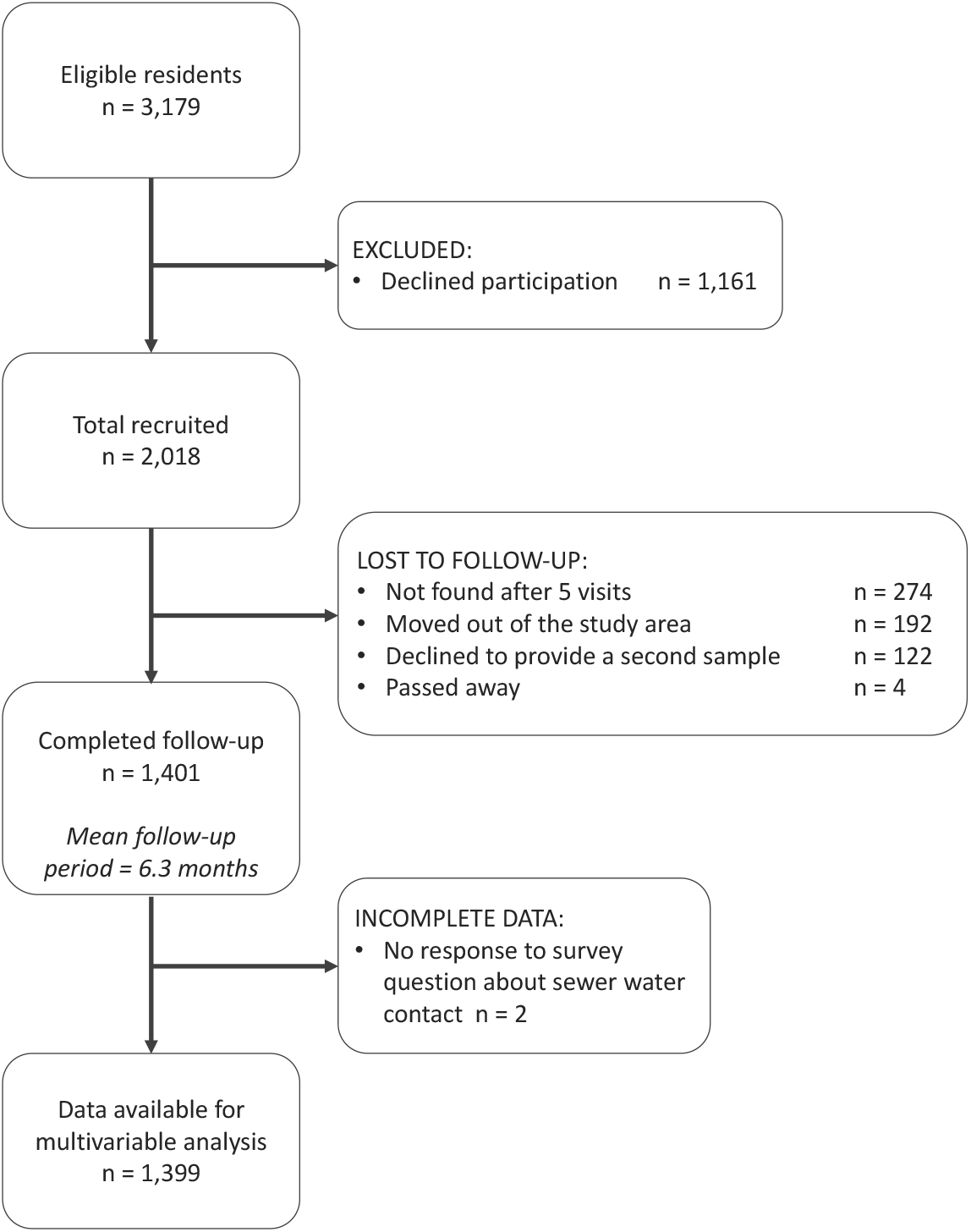
The study participant flow chart in line with the STROBE (Strengthening the Reporting of Observational Studies in Epidemiology) statement (http://www.strobestatement.org))

Between the two serosurveys there was serological evidence of 72 leptospiral infections in the cohort, with an overall infection rate of 51.4 (95%CI 40.4, 64.2) infections per 1,000 follow-up events. Valleys 2 and 3 had high estimated infection rates with 66.4 (95%CI 47.3, 90.2) infections per 1,000 follow-up events and 49.6 (95%CI 33.6, 69.9) infections per 1,000 follow-up events, respectively, compared to 23.2 (95%CI 9.2, 46.9) infections per 1,000 follow-up events in Valley 1. In the rat ecology study: a rat was captured in 129 (9.0%) out of 1,512 trapping-days; 263 (37.4%) out of 703 track plate days had at least one positive plate; 28.5%, 19.7% and 25.9% of the 580 sampled locations found at least one sign of active burrows, faecal droppings and trails, respectively.

### 3.2 Exploratory analysis and model selection

The results from the exploratory multivariable analysis of rattiness are shown in Table 1. The linear splines used were informed by the functional forms shown in Supplementary file 1. The relationship between rattiness and relative elevation demonstrates a trade-off between the high availability of food sources at the bottom of the valley and high risk of flooding which prevents the establishment of burrows. In the lowest elevation areas (0-8m above the bottom of the valley), relative elevation and rattiness were positively associated with an increase of 0.04 (95%CI 0.00, 0.07) rattiness units per 1m; when interpreting the magnitude of effect estimates note that, by definition, rattiness is defined so as to have variance one. Rattiness then peaked at an elevation of 8m before declining with increasing elevation by 0.04 (95%CI -0.09, 0.01) units until an elevation of 22m. Rattiness started to increase again above this elevation by 0.06 (95%CI 0.00, 0.10) units per metre. Rattiness decreased with increasing distance from large refuse piles, a source of food and harbourage, by 0.07 (95%CI -0.13, -0.01) units per 10m distance until a distance of 50m, beyond which there was little effect. Impervious land cover (defined as the proportion of the area within a 30m radius around each sampling location classified as pavement or building) was negatively associated with rattiness, decreasing by -0.05 (95%CI -0.08, -0.01) units for every 10% increase in impervious cover.

**Table 1:** Multivariable linear regression analysis of predictors for rattiness (note that rattiness is a unit-variance random variable when interpreting the magnitude of effect estimates)

| Variable | Estimate (95%CI)<sup>1</sup> |
| --- | --- |
| Relative elevation (per 1m increase)<sup>2</sup> |  |
| 0-8m | 0.04 (0.00, 0.07) |
| 8-22m | -0.04 (-0.09, 0.01) |
| >22m | 0.06 (0.00, 0.10) |
| Distance to large refuse piles (per 10m increase)<sup>2</sup> |  |
| 0-50m | -0.07 (-0.13, -0.01) |
| >50m | 0.02 (-0.05, 0.09) |
| Impervious land cover (per 10% increase) | -0.05 (-0.08, -0.01) |

<sup>1</sup> CI, Confidence interval;
<sup>2</sup> The effects of relative elevation and distance to refuse are modelled as broken linear models with transitions at 8m and 22m, and 50m, respectively. This was informed by the relationship described by Generalized Additive Modelling (Supplementary file 1)

In the community cohort data, the univariable analysis identified several risk factors that increased a resident’s risk of leptospiral infection (Table 2). Variables with statistically significant associations (at the conventional 5% level) with infection risk were identified in two of the four domains: demographic and social status and behavioural exposures. Within the demographic and social status domain, risk of infection increased with age and was found to be higher for male participants and those living in Valleys 2 and 3. In the behavioural exposures domain, participants who had had frequent contact with floodwater in the last six months were more likely to be infected. Two individuals were excluded from the multivariable analysis (n=1,399) because of missing data for the floodwater exposure survey question.

**Table 2:**
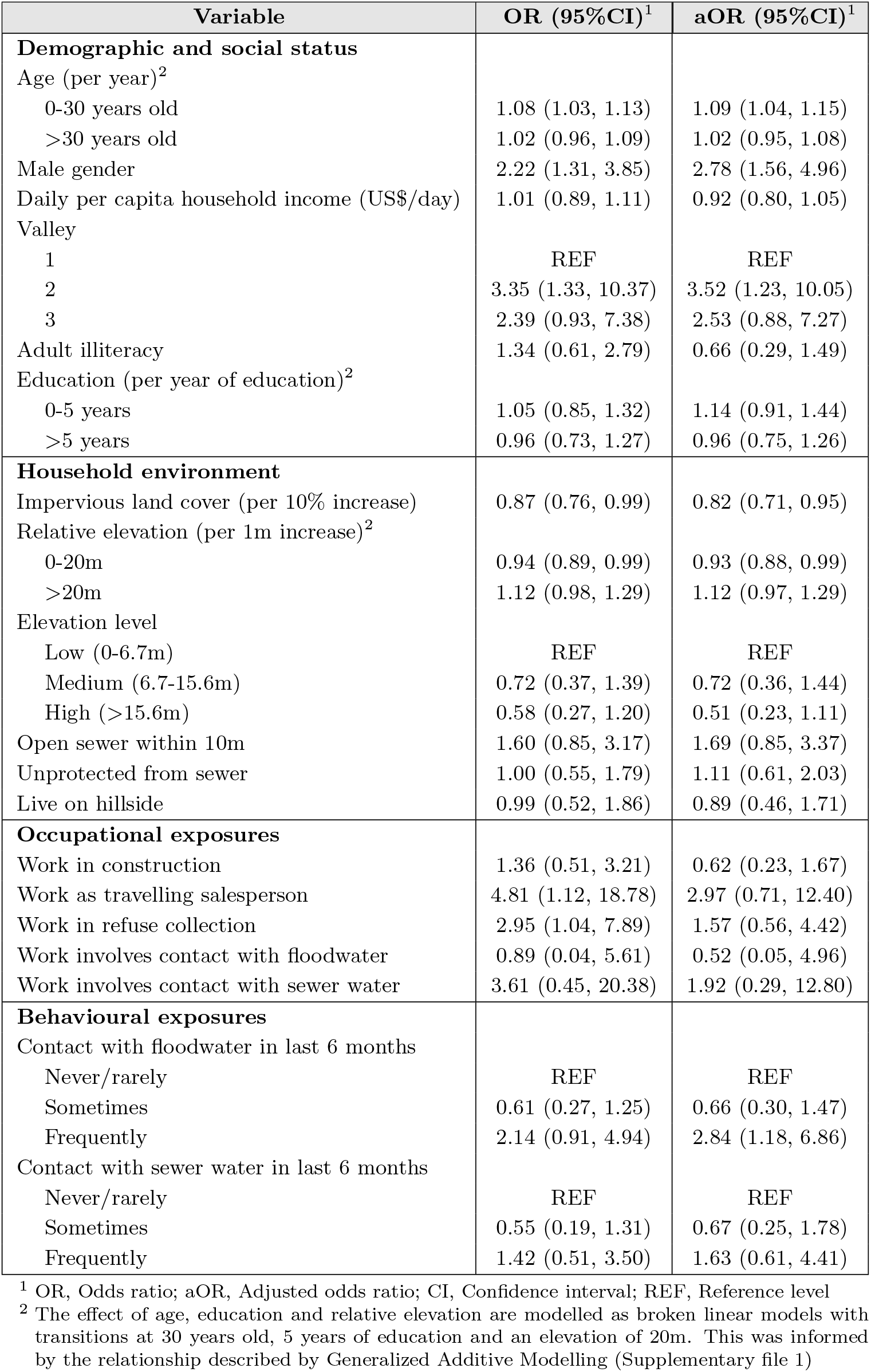
Univariable mixed effects logistic regression analysis of human risk factors for leptospiral infection

In the exploratory results from the multivariable model there was strong evidence of an interaction between rattiness and household elevation level on human infection risk (see Supplementary file 3 for all parameter estimates for this model). At the high elevation level area, a unit increase in mean predicted rattiness at the household location was estimated to increase the odds of infection by 6.92 (95%CI 1.88, 25.47). In contrast, in the low and medium elevation areas there was no evidence of a relationship between rattiness and infection risk, as shown in Figure 4. Consequently, this interaction effect was also included in the rattiness-infection joint model.

**Figure 4:**
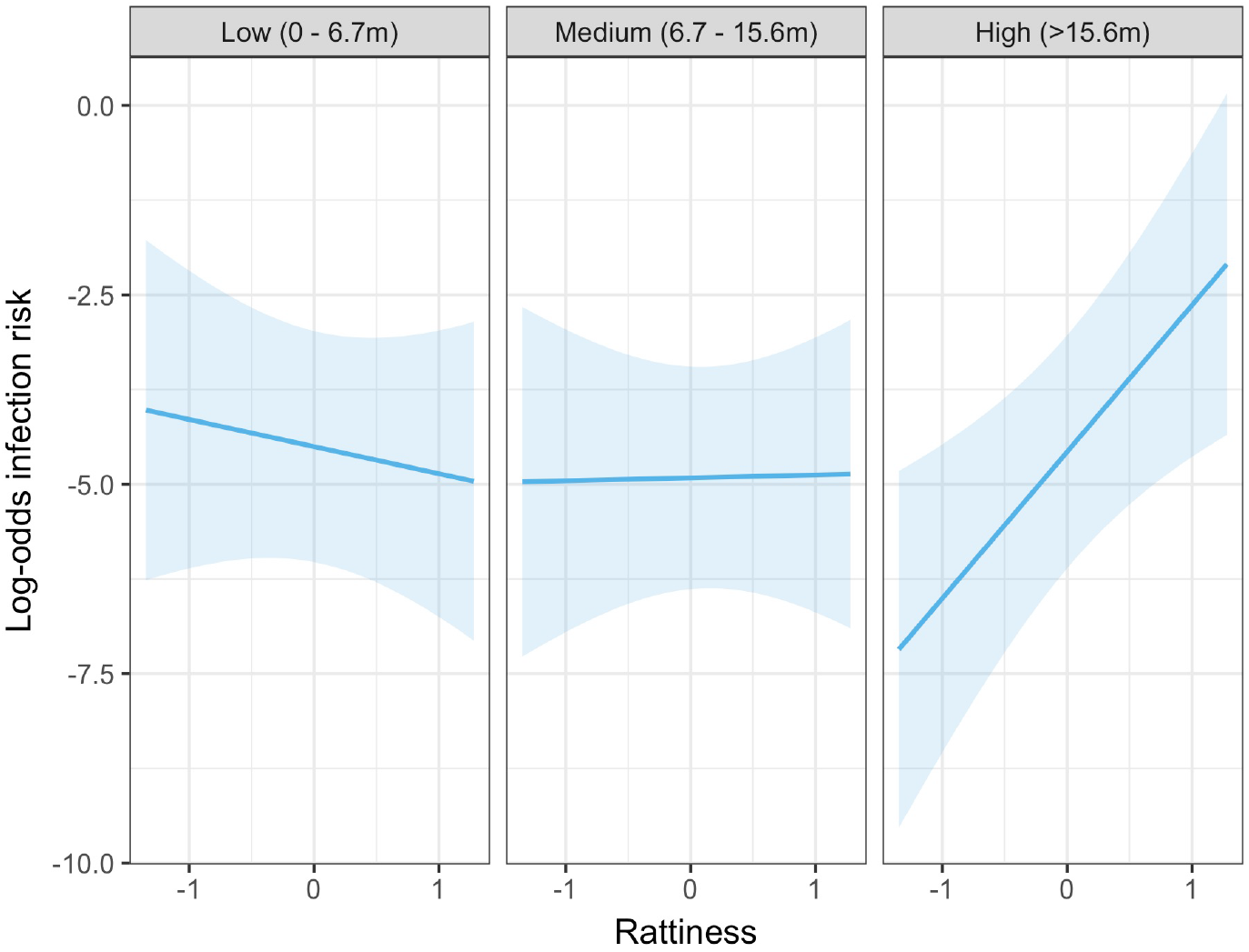
Predicted relationship between rattiness and infection risk from the multivariable mixed effects logistic regression demonstrating evidence of an interaction with elevation level (low, medium and high). Shown on the log-odds scale with shaded areas corresponding to 95% confidence intervals.

The explanatory variables selected in the rat and human multivariable analyses were then entered into the full rattiness-infection joint model with the functional forms included in Table 1. To test for residual spatial correlation in the human infection data after controlling for explanatory variables and rattiness, we fitted the joint model with an additional spatial Gaussian process in the human infection linear predictor. The estimated value for the scale of spatial correlation for this Gaussian process was less than 1m and indistinguishable from household-level variation. We consequently fitted the joint model specified in Equation (3) which assumes that there is no residual spatial correlation in the human infection data.

### 3.3 Joint rattiness-infection model

Human infection risk factors, rattiness predictors and other model parameters estimated using the joint rattiness-infection model are shown in Table 3. Infection risk was strongly associated with age, with an individual experiencing an increased odds of infection of 1.09 (95%CI 1.04, 1.19) for every year of life up until 30 years of age, and 1.02 (95%CI 0.92, 1.09) for each additional year thereafter. Male participants were more likely to be infected than female participants (OR 2.69 95%CI 1.58, 5.89). Compared with individuals living in Valley 1, those living in Valley 2 had a higher estimated odds of infection (OR 2.91 95%CI 1.03, 20.82). Individuals living in the medium (OR 0.77 95%CI 0.31, 1.66) and high (OR 0.67 95%CI 0.11, 1.64) elevation areas had a lower estimated odds of infection relative to those living in the low elevation area where the open sewers are and flooding risk is higher, but these differences were not significant at the conventional 5% level.

**Table 3:** Parameter estimates for the full joint rattiness-infection model

| Parameter | Estimate (95%CI) |
| --- | --- |
| **Human infection risk factors** | **OR** |
| Age (per year) |  |
| 0-30 years old | 1.09 (1.04, 1.19) |
| >30 years old | 1.02 (0.92, 1.09) |
| Male gender | 2.69 (1.58, 5.89) |
| Daily per capita household income (US\$/day) | 0.93 (0.74, 1.05) |
| Valley |  |
| 1 | REF |
| 2 | 2.91 (1.03, 20.82) |
| 3 | 2.28 (0.86, 14.00) |
| Elevation level |  |
| Low (0-6.7m) | REF |
| Medium (6.7-15.6m) | 0.77 (0.31, 1.66) |
| High (>15.6m) | 0.67 (0.11, 1.64) |
| Work as travelling salesperson | 3.16 (0.38, 20.57) |
| Contact with floodwater in last 6 months |  |
| Never/rarely | REF |
| Sometimes | 0.62 (0.18, 1.39) |
| Frequently | 2.47 (0.67, 7.41) |
| *Rattiness* (per unit rattiness) |  |
| $\xi_{low}$ | 1.14 (1.05, 1.53) |
| $\xi_{med}$ | 1.25 (1.08, 1.74) |
| $\xi_{high}$ | 3.27 (1.68, 19.07) |
| $\sigma^2$ (variance of household-level random effect) | 1.36 (0.23, 5.35) |
| **Rattiness variables** |  |
| Relative elevation (per 1m increase)<sup>2</sup> |  |
| 0-8m | 0.05 (-0.01, 0.13) |
| 8-22m | -0.06 (-0.16, 0.02) |
| >22m | 0.05 (-0.03, 0.14) |
| Distance to large refuse piles (per 10m increase)<sup>3</sup> |  |
| 0-50m | -0.10 (-0.21, 0.02) |
| >50m | 0.03 (-0.11, 0.17) |
| Impervious land cover (per 10% increase) | -0.07 (-0.14, -0.01) |
| **Rattiness parameters** |  |
| $\alpha_{traps}$ | -2.94 (-3.27, -2.65) |
| $\alpha_{plates}$ | -2.06 (-2.50, -1.74) |
| $\alpha_{burrows}$ | -1.41 (-1.67, -1.16) |
| $\alpha_{faeces}$ | -2.82 (-3.83, -2.32) |
| $\alpha_{trails}$ | -2.22 (-2.96, -1.76) |
| $\sigma_{traps}$ | 0.72 (0.45, 0.97) |
| $\sigma_{plates}$ | 2.37 (2.05, 2.68) |
| $\sigma_{burrows}$ | 1.28 (1.08, 1.45) |
| $\sigma_{faeces}$ | 2.36 (1.80, 3.34) |
| $\sigma_{trails}$ | 2.43 (1.85, 3.12) |
| $\psi$ | 0.67 (0.29, 1.00) |
| $\phi$ | 9.23 (3.21, 18.24) |

Infection risk was positively associated with rattiness for households situated in all three elevation levels. However, while the effect size (per unit increase in rattiness) was similar in the low (OR 1.14 95%CI 1.05, 1.53) and medium (OR 1.25 95%CI 1.08, 1.74) elevation areas, in the high elevation area the effect of increasing rattiness on infection risk was significantly stronger (OR 3.27 95%CI 1.68, 19.07). This interaction effect between rattiness and household elevation level on human infection risk was confirmed with a test for evidence against the null hypothesis that *ξ*_*low*_ = *ξ*_*med*_ = *ξ*_*high*_ (*p* = 0.026, χ^2^ = 7.33, *df* = 2).

Parameter estimates for the rattiness variables were very similar to the estimates from the exploratory linear regression (Table 1), with a slightly higher effect size for the distance to refuse piles and land cover variables. There was evidence of small-scale spatial correlation in rattiness (*ϕ*= 9.23*m* 95%CI 3.21, 18.24m) corresponding to a spatial correlation range (the distance at which the correlation reduces to 5%) of approximately 28m. The estimate for *ψ* of about 0.67 (95%CI 0.29, 1.00) indicates that the majority of the unexplained variation in rattiness is spatially structured, with the remainder modelled as a nugget effect.

### 3.4 Spatial prediction

There was heterogeneous spatial variation in predicted rattiness. The numerous small regions of high rattiness in Figure 5A are indicative of the small-scale spatial correlation in the data. The low elevation areas in the central length of each valley (relative elevation is shown in Figure 5E with the contours marking the low, medium and high elevation areas) had high mean predicted rattiness. High rattiness was also predicted in several high elevation areas, for example the northern tip of Valley 3 and several small hotspots along the three valley’s high elevation sides.

**Figure 5:**
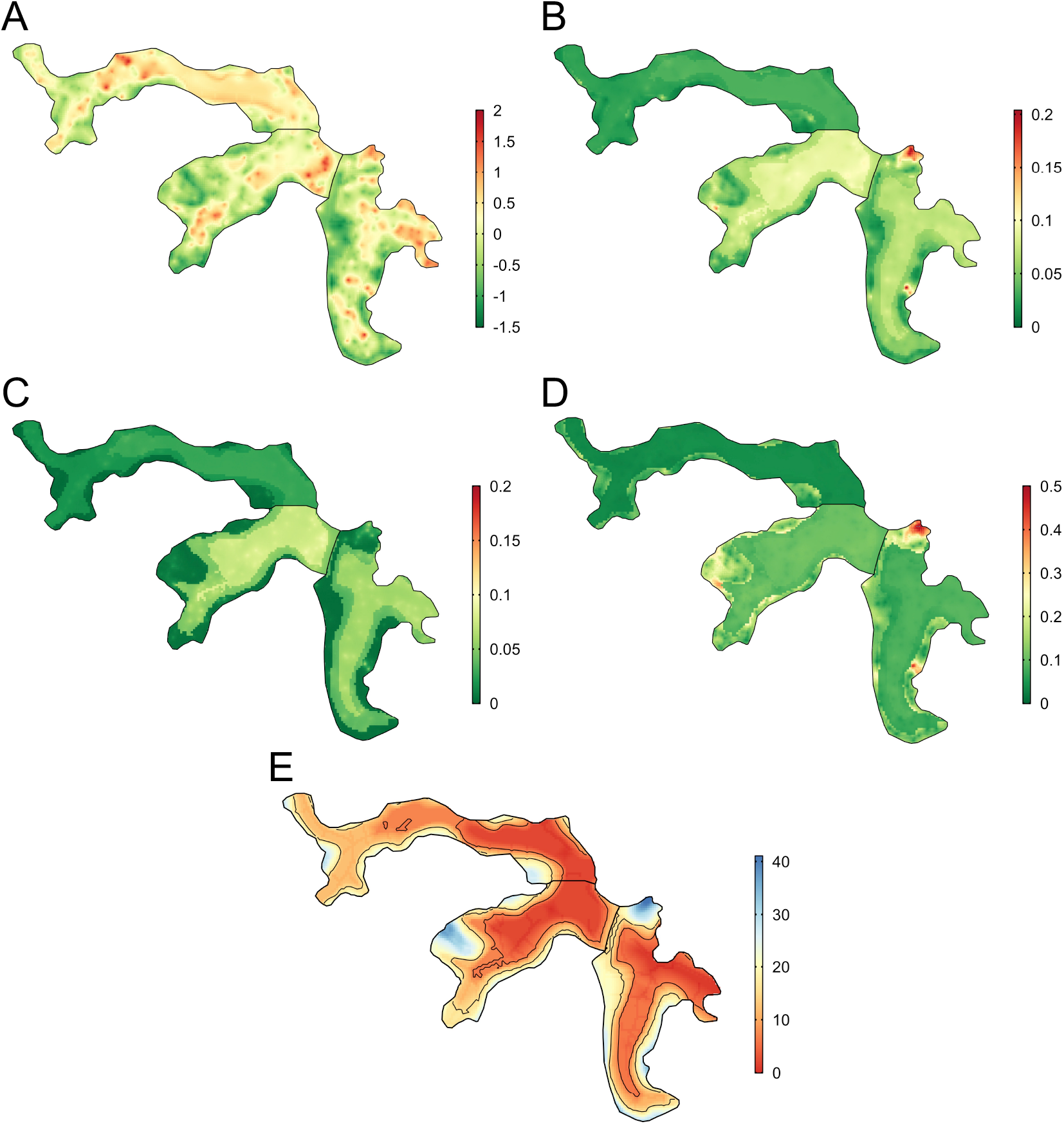
Joint rattiness-infection model predictions. A) Mean predicted rattiness; B) Mean predicted leptospiral infection risk for 30-year-old male participants with a household per capita income of USD$1/day who never/rarely have contact with floodwater and do not work as a travelling salesperson; C) lower 95% prediction interval for predicted infection risk; D) upper 95% prediction interval for predicted infection risk; E) Elevation (metres) relative to the bottom of valley (low, medium and high levels are shown with contours)

To illustrate the spatial variation in infection risk within the study area, prediction maps are shown in Figure 5B for a 30-year-old male participant with a household per capita income of USD$1/day who never or rarely had contact with floodwater in the previous six months and did not work as a travelling salesperson. Infection risk was low across most of Valley 1 (*<*2.5%), with marginally higher average values found in the central low elevation area (2.5-5%). Risk was consistently higher across most of Valleys 2 and 3 (7.5-15%), with the effect of elevation on risk clearly visible. In areas with higher and more spatially heterogeneous predicted infection risk, for example in the central region of Valley 2, this was driven by high levels of predicted rattiness. The stronger estimated effect of rattiness on infection risk in higher elevation areas was particularly visible in Valleys 2 and 3, as seen in the three hotspots with risk reaching 20% and the moderate risk hotspots along the sides of both valleys. Prediction intervals were relatively narrow across most of the study area (Figures 5C and 5D) with greater uncertainty in the high risk areas.

## 4 Discussion

We developed and applied a novel framework for joint spatial modelling of disease reservoir abundance and human infection risk to a community-based cohort study and fine-scale rat ecology study. We found that higher levels of rattiness, our proxy for rat abundance, at the household location were associated with a higher risk of leptospiral infection for residents across the entire study area. Importantly, we found that a unit increase in rattiness in high elevation areas was associated with an almost three times higher odds ratio for infection than in low and medium elevation areas. To our knowledge this is the first study to jointly model rodent abundance and human infection data for a rodent-borne zoonosis. The findings provide new insights into how the dominant mechanisms of *Leptospira* transmission within complex urban settings may vary over small distances, as a result of interactions between rats, the environment, geography, and local epidemiology.

The finding that rattiness was associated with infection risk indicates that the spatial distribution of rat populations was an important driver of transmission close to the household across the entire study area. This is consistent with a recent study investigating the predictive power of household rat infestation scores for human infection [30]. There was no residual spatial correlation in the infection data after accounting for rattiness in our analysis, possibly suggesting that previously unexplained spatial heterogeneity in risk could be driven by variation in rattiness [12]. Our model also predicted high average rattiness across the low elevation areas where leptospiral transmission is high [10, 12]. This supports the hypothesis that abundant rat populations are responsible for high levels of observed environmental contamination across these lower areas [21, 60], and consequently increased infection risk.

The identified interaction between elevation and rattiness on infection risk suggests that relatively small changes in environment and topography can modify transmission pathways within an urban community. The weaker effect of rattiness on infection risk at low and medium elevation areas relative to high elevation areas may be explained by differences in their hydrological profiles. While high rat abundance in low and medium areas results in high leptospiral contamination, these areas are prone to high levels of water runoff and flooding. This disperses the pathogen across low elevation areas. The ability of leptospires to persist in the environment for weeks or months means that this process can significantly increase environmental risk in low elevation areas for long periods. This process disconnects shedding and infection events in space and time [1] and obscures the relationship between infection risk and rattiness in low and medium elevation households.

In contrast, high elevation areas have lower levels of water runoff and flooding due to improved drainage and sewage systems, and a smaller upstream catchment area for rainfall. Leptospires are consequently less likely to be washed away from the location at which they were shed, and environmental risk remains more localised and strongly associated with the spatial distribution of rats. This hypothesised role of hydrology in the aggregation and dispersal of leptospires [1] is supported by a recent study in low elevation areas of Pau da Lima which found that soil contamination was not associated with local rat activity [20]. However, our finding that rattiness was associated with infection in low and medium elevation areas suggests that the spatial distribution of environmental risk in these areas is not entirely determined by water dispersal.

Interestingly, a previous study of surface waters in Pau da Lima found that the probability of a sample being positive for *Leptospira* was highest in low elevation areas and lowest in medium elevation areas, with no significant difference between low and high elevation areas [21]. This is consistent with our findings and suggests that there may be a ‘washing out’ of locally deposited leptospires in medium elevation areas but not in high elevation areas.

This has several implications for disease control strategies which aim to reduce environmental risk. Improving drainage systems at all elevation levels can reduce the dispersal of leptospires from high to low elevation areas. Closure of sewer systems, which generally run through low elevation areas, can protect local residents from exposure and reduce the introduction of additional contamination from upstream sewer water. Paving over soil surfaces can reduce the surface area over which leptospires can persist [6], reducing environmental risk further.

A reduction in the dispersal and accumulation of bacteria will result in more localised environmental risk, as was observed in the high elevation areas in this study. Higher risk will then be found in areas with a high abundance of infected rats. This may also reduce environmental exposure for rats, thereby lowering shedding rates and acting as a feedback loop into the *Leptospira* transmission cycle [61]. Given the limited and short-term impact of chemical rodenticide campaigns on Norway rat abundance in these settings [62], longer-term environment management strategies targeted at rattiness hotspots may also be needed to reduce the availability of key predictors of rattiness, such as large refuse piles and vegetation and soil land cover. Funding and political will for large-scale infrastructural interventions is often limited in marginalised urban settings and small-scale community-based interventions which target these mechanisms should be evaluated.

Transmission is dynamic in space and time and the alignment of conditions which enable spillover infection can vary over time [1]. Our study was designed to explore the spatial variation in rattiness and infection risk in Pau da Lima during the driest period of the year, and it may not be representative of transmission mechanisms during the rainy season. There is some evidence, however, that this may not necessarily be the case, with two recent studies in Pau da Lima reporting low seasonal variation in both rat abundance [63] and spatial infection risk patterns [12]. Nonetheless, future studies across different time periods are needed to establish the role of rat abundance in *Leptospira* transmission.

In this study we used household location to link rattiness to an individual, under the assumption that the majority of their exposure occurs close to home. Given the spatially heterogeneous distribution of rattiness and environmental contamination [21] within the community, future epidemiological studies of leptospirosis and zoonotic spillover could benefit from trying to pinpoint key sources of infection away from the household using GPS mobility data, as has been attempted in a small study previously [64]. The rattiness-infection framework could then be extended to model cumulative environmental exposure to the rattiness surface by integrating along a person’s trajectory as they move around the community.

Our framework did not account for disease dynamics within rat populations. Given that 80% of rats are estimated to be actively shedding *Leptospira* in Pau da Lima [27, 28] and prevalence in rats is generally high in urban areas globally [22–26], the use of rattiness as a proxy for rat shedding appears reasonable and it may be a useful proxy in other epidemiological studies. However, in other zoonotic spillover systems where pathogen release does not occur at a high and homogeneous rate across the reservoir host population this may not be the case and accounting for spatially heterogeneous or time-varying [65] disease dynamics will be important.

The rattiness-infection modelling framework is a flexible tool for exploring the spatial association between reservoir abundance, the environment and human health outcomes. It provides a statistically principled method for joint spatial modelling of infection risk and multiple indices of reservoir abundance, pooling data between indices and directly accounting for uncertainty in their measurement in all parameter estimates and predictions. The framework’s geostatistical structure includes spatially continuous predictors for abundance and accounts for spatial correlation, enabling mapping of both infection risk and rattiness. This can be useful for identifying high-risk areas and targeting control. One inherent limitation is its dependence on the availability of spatially continuous environmental variables and abundance data, both of which are prone to high measurement error. This can result in high uncertainty in the model parameter estimates and predictions, as demonstrated by the wider confidence intervals for risk factors in the joint model compared to the standard mixed-effects logistic regression analysis. An additional benefit of the geostatistical structure is that abundance measurements do not have to be taken at the household location, providing some flexibility in the design of eco-epidemiological studies and indices used.

In conclusion, we have developed a framework that may have broad applications in delineating complex animal-environment-human interactions during zoonotic spillover and identifying opportunities for public health intervention. We demonstrate its potential by applying it to *Leptospira* in an urban setting, finding evidence that the extent to which local rat shedding drives spillover transmission is moderated by elevation, most likely a proxy for water runoff. Future work examining these transmission mechanisms in similar settings and across different time points will be key to establishing how generalisable these results are.

## Data Availability

Rat and human data analysed in this study have been deposited in OSF (https://doi.org/10.17605/OSF.IO/AQZ2Y). However, the human data has been anonymised and household coordinates and valley ID have been removed. Modelling functions, R scripts and metadata for analyses in this manuscript are publicly available at https://github.com/maxeyre/Rattiness-infection-framework.

## CRediT authorship contribution statement

**Max T. Eyre**: Conceptualization, Methodology, Software, Validation, Formal analysis, Investigation, Data curation, Visualization, Writing-original draft, Writing - Review and editing.

**Fábio N. Souza, Ticiana S. A. Carvalho-Pereira, Nivison Nery Jr**., **Daiana de Oliveira, Jaqueline S Cruz, Kathryn P. Hacker, Gielson A. Sacramento** and **Elsio A. Wunder Jr**: Investigation, Data curation, Writing - Review and editing.

**Hussein Khalil** and **James E. Childs**: Writing - Review and editing.

**Mitermayer G. Reis**: Project administration, Conceptualization, Investigation, Resources, Supervision, Review and editing, Funding acquisition.

**Mike Begon**: Conceptualization, Supervision, Writing - Review and editing.

**Albert I. Ko**: Conceptualization, Funding acquisition, Writing - Review and editing.

**Peter J. Diggle** and **Emanuele Giorgi**: Conceptualization, Methodology, Software, Supervision, Writing - Review and editing.

**Federico Costa**: Project administration, Conceptualization, Investigation, Resources, Supervision, Writing - Review and editing, Funding acquisition.

## Acknowledgements

We thank the residents and community leaders of Pau da Lima community for their support and participation in this study. This work was supported by the Oswaldo Cruz Foundation and Secretariat of Health Surveillance, Brazilian Ministry of Health, the National Institutes of Health of the United States (grant numbers F31 AI114245, R01 AI052473, U01 AI088752, R01 TW009504 and R25 TW009338); the Wellcome Trust (102330/Z/13/Z), and by the Fundação de Amparo á Pesquisa do Estado da Bahia (FAPESB/JCB0020/2016). M.T.E is supported by an MRC doctorate studentship. F.N.S. participated in this study under a FAPESB doctorate scholarship.

## Supplementary file 1 - Functional form of continuous explanatory variables

**Figure S1:**
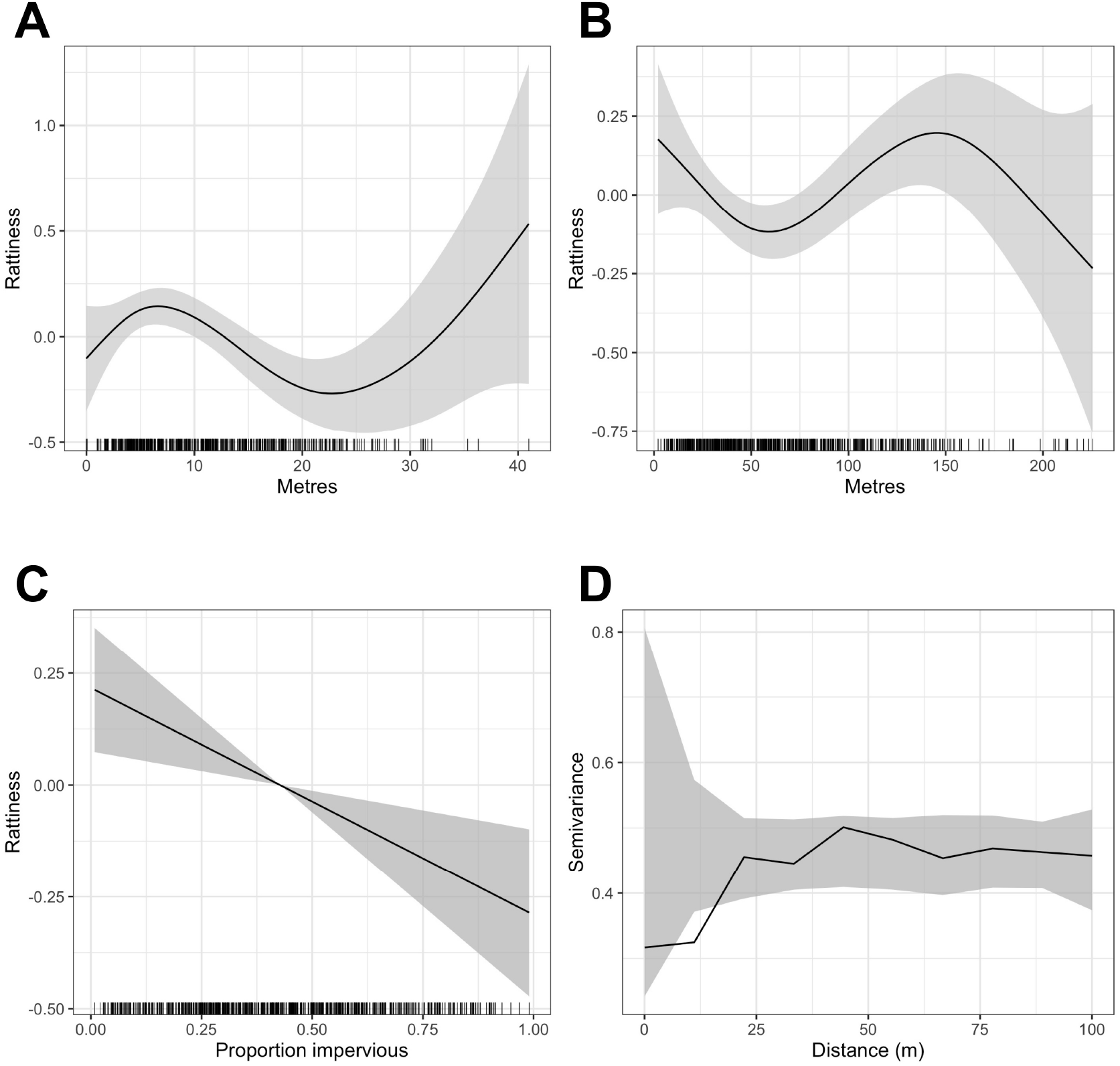
Generalized Additive Model (GAM) partial dependence plots for the unstructured random variation in rattiness, *Û*_*i*_, plotted against the continuous explanatory variables considered in the analysis (shaded areas correspond to 95% confidence intervals): A) elevation relative to the bottom of valley, B) distance to large refuse piles, C) impervious land cover in 20m radius buffer around sampling point. *Û*_*i*_ are estimated using a non-spatial model which excludes all covariates. Panel D) is a variogram computed from *Û*_*i*_ using a non-spatial model that includes all of the covariates; the dashed lines correspond to 95% confidence intervals under the assumption of spatial independence.

**Figure S2:**
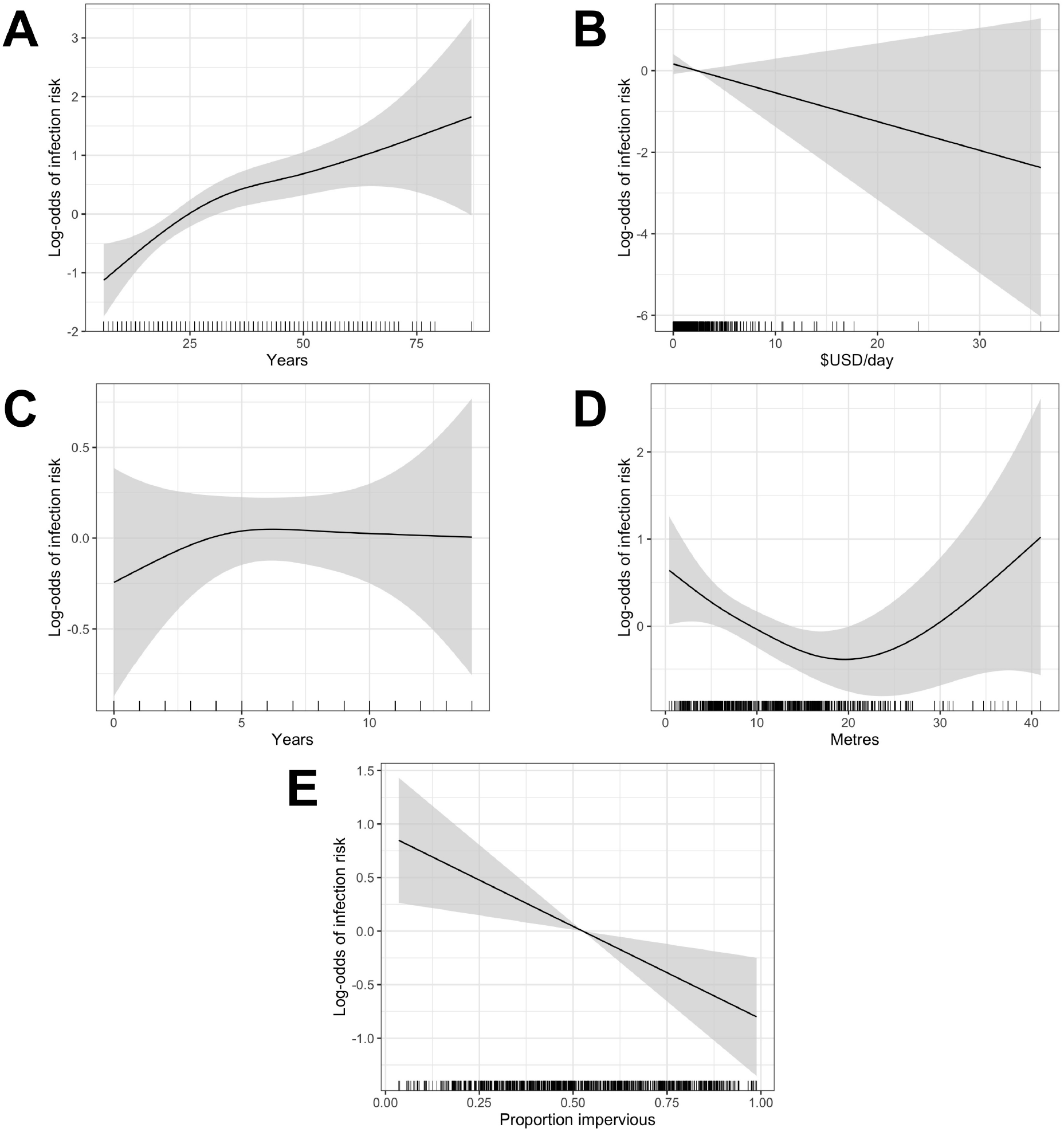
Generalized Additive Model (GAM) partial dependence plots for human infection risk plotted against the continuous explanatory variables considered in this analysis (shaded areas correspond to 95% confidence intervals): A) age, B) household per capita income (in USD), C) years of education, D) household elevation relative to the bottom of valley, E) impervious land cover in 20m radius buffer around household.

